# A HOME-TREATMENT ALGORITHM BASED ON ANTI-INFLAMMATORY DRUGS TO PREVENT HOSPITALIZATION OF PATIENTS WITH EARLY COVID-19: *A MATCHED-COHORT STUDY (COVER 2)*

**DOI:** 10.1101/2021.09.29.21264298

**Authors:** Elena Consolaro, Fredy Suter, Nadia Rubis, Stefania Pedroni, Chiara Moroni, Elena Pastò, Maria Vittoria Paganini, Grazia Pravettoni, Umberto Cantarelli, Norberto Perico, Annalisa Perna, Tobia Peracchi, Piero Ruggenenti, Giuseppe Remuzzi

## Abstract

**Background and Aim:** While considerable success has been achieved in the management of patients hospitalized with severe coronavirus disease 2019 (COVID-19), far less progress has been made with early outpatient treatment. We assessed whether the implementation of a home treatment algorithm – designed based upon on a pathophysiologic and pharmacologic rationale - during the initial, mild phase of COVID-19, could effectively reduce hospital admissions.

**Methods:** This fully academic, matched-cohort study evaluated outcomes in 108 consecutive consenting patients with mild COVID-19 managed at home by their family doctors from January 2021 to May 2021, according to the proposed treatment algorithm and in 108 age-, sex-, and comorbidities-matched patients who were given other therapeutic schedules (ClinicalTrials.gov: NCT04854824). The primary outcome was COVID-19-related hospitalization. Analyses were by intention-to-treat.

**Results:** One (0.9%) patient in the ‘recommended’ cohort and 12 (11.1%) in the ‘control’ cohort were admitted to hospital (P=0.0136). The proposed algorithm reduced, by 85%, the cumulative length of hospital stays (from 141 to 19 days) and related costs (from € 60.316 to € 9.058). Only 9.8 patients needed to be treated with the recommended algorithm to prevent one hospitalization event. The rate of resolution of major symptoms was numerically, but not significantly, higher in the ‘recommended’ compared to the ‘control’ cohort (97.2% versus 93.5%, respectively; P=0.322). Other symptoms lingered in a lower proportion of patients in the ‘recommended’ than in the‘control’ cohort (20.4% versus 63.9%, respectively; P<0.001), and for a shorter period.

**Conclusion:** The adoption of the proposed outpatient treatment algorithm during the early, mild phase of COVID-19 reduced the incidence of subsequent hospitalization and related costs.

## 1. Introduction

Over the past two years the novel coronavirus SARS-CoV-2 (Severe Acute Respiratory Syndrome Coronavirus 2), which causes coronavirus disease 2019 (COVID-19), has quickly spread globally, reaching pandemic proportions (1). Through genetic evolution resulting in multiple variants (2), SARS-CoV-2 has been responsible for several pandemic waves worldwide (1). The clinical manifestations of COVID-19 disease are broad, spanning asymptomatic infection, mild upper respiratory tract and/or mild extrapulmonary symptoms, and including severe pneumonia, acute respiratory distress syndrome and multiorgan system dysfunction, and even death (3,4). During the early phase of COVID-19 when patients are at home they are usually not seriously ill with acute respiratory distress, but present a variety of initially mild/moderate symptoms, including fever, cough, tiredness, shortness of breath and chills, a sore throat, headache, musculoskeletal pain, and a new loss of taste and smell (5).

While drug/biological treatment options for severely ill COVID-19 patients requiring hospitalization are now available (6–11), interventions that can be administered by primary care physicians at home have been more difficult to determine and controversial (12). Nonetheless, the early initiation of treatment for COVID-19 might improve clinical outcomes, providing a potential window for immediate benefits by intervening before the development of severe disease, and possibly limiting or preventing the risk of patient hospitalization.

Although guidelines or recommendations for managing patients with suspected or confirmed COVID-19 in the community have recently been made available by national health authorities (13,14), most family doctors initially treated their patients with various treatment regimens they believed appropriate based on their clinical expertise. Based on the increasing available knowledge on the pathophysiology underlying the mild/moderate symptoms encountered at the onset of the illness (15,16), we recently published a proposed regimen of simple drugs that should theoretically better fit these mechanisms (17). The proposed treatment recommendation (17) is based on three pillars: i) intervene at the very onset of mild/moderate symptoms at home; ii) start therapy as early as possible after the family doctor has been contacted by the patient, without awaiting the results of a nasopharyngeal swab; iii) rely on non-steroidal anti-inflammatory drugs, especially relatively selective cyclooxygenase-2 (COX-2) inhibitors (18,19), an approach intended to limit excessive host inflammatory responses to viral infection (16,17).

In a recent academic matched-cohort study (20), we found that early treatment of COVID-19 patients at home by their family doctors, according to the proposed recommendation regimen, almost completely prevented the need for hospital admission due to progression toward more severe illness (2 out of 90 patients), compared to patients in the ‘control’ cohort, who were treated at home according to their family physician’s assessments (13 out of 90 patients). However, the rate of hospitalization was a secondary outcome of the study, and the possibility that this is a random finding cannot be excluded definitely. Thus, we considered the observed reduction in patient hospitalizations a hypothesis-generating finding that provides the background for the present, new matched-cohort study. The primary aim of this study was to test the effect of COVID-19 treatment at home on this outcome, according to the proposed recommendation algorithm.

## 2. Materials and methods

### 2.1 Study design and participants

This in an observational study that involved two matched cohorts of COVID-19 patients. The ‘recommended algorithm’ cohort included 108 patients treated at home by their family physicians who expressed interest in participating in the study and followed the published proposed treatment recommendation (see Supplementary Methods) (17). They were from the Varese, Bergamo, and Teramo provinces (Italy) and prospectively engaged their patients between January and May 2021. These family doctors applied the recommended algorithm at the onset of symptoms, or within a few days of being contacted by patients. The physicians provided patients with detailed information about the objectives and design of study and collected signed consent forms. They were asked to complete an online questionnaire with information on the outcomes of COVID-19 symptoms/illness that are relevant to addressing the primary, secondary and safety aims of the study. The Istituto di Ricerche Farmacologiche Mario Negri IRCCS (Bergamo, Italy) was the coordinator of the project, promoted through online institutional media. Male and female adults, aged ≥ 18 years, with early mild symptoms of COVID-19, who started the recommended treatment without awaiting the results of a nasopharyngeal swab, if any, were eligible to participate in the study.

As a control cohort, 108 historic COVID-19 patients were retrospectively considered. These patients had been enrolled in the “Study of the Genetic Factors that Influence the Susceptibility to and Severity of COVID-19” (the ORIGIN study, conducted by the Istituto di Ricerche Farmacologiche Mario Negri, IRCCS (ClinicalTrials.gov; NCT04799834), and treated at home by their family doctors with drug regimens not necessarily guided by those proposed in the recommendation algorithm. They were matched by age, sex, comorbidities (hypertension, diabetes, cardiovascular diseases, overweight, chronic kidney disease) with patients in the ‘recommended algorithm’ cohort. Notably, the ORIGIN study collects, among other things, all clinical information intended for the analyses of the ‘recommended algorithm’ cohort from the population of COVID-19 patients living in the Bergamo province.

In both cohorts, subjects who required immediate hospitalization, according to the family physician’s assessment, because of severe COVID-19 symptoms at onset, were excluded.

### 2.2 Outcome variables

The primary outcome was the proportion of patients requiring hospitalization due to clinical worsening of the illness in the two treatment cohorts.

Secondary outcomes included: i) Compliance to the algorithm in the cohort that adopted the proposed treatment recommendations, defined as adherence to recommended schedule of treatment; ii) Number of days between onset of symptoms and the start of anti-inflammatory therapy; iii) The proportion of patients in the two cohorts with complete resolution of major symptoms (‘complete remission’) defined as recovery from these symptoms, namely no fever, SpO_2_ >94% and/or no dyspnea, cough, rhinitis, pain (myalgia, arthralgia, chest pain, headache, sore throat), vertigo, nausea, vomiting or diarrhea, sicca syndrome or red eyes; iv) The proportion of patients in the two cohorts with persistent other symptoms, such as anosmia, ageusia/dysgeusia, lack of appetite, fatigue. In addition, the duration of persistence of these symptoms (<30 days, or 30 to 60, or >60 days) was evaluated; v) Time (in days) spent in an intensive care unit, sub-intensive care unit, and ordinary units by patients who required hospital admission in the two cohorts; vi) Cumulative hospitalization costs (in euro) for patients admitted to hospital in the two cohorts. Potential baseline confounders such as age, sex, and concomitant diseases that could increase the risk of severe COVID-19 illness were predefined (21–23). Moreover, serious (SAE) and non-serious adverse events (AE) related to the administered treatments according to recommendations were assessed. The severity/non-severity of the observed events and their causal relationships with treatments were determined by the family doctor in charge of the patients.

### 2.3 Samples size and statistical analysis

Based on our recent findings (20), we assumed that the proportion of hospital admissions in the ‘historic control’ cohort, when patients were treated by their family doctors according to drug regimens not necessarily guided by those proposed in our recommendation algorithm, is 0.1444, and that in the ‘recommended algorithm’ cohort it is 0.0222. Based on the above assumption, a sample size of 85 patients per group (170 total) would achieve 80% power to detect a difference between the group proportions of 0.1222 (two-sided log rank test, alpha=0.05). Assuming a 20% drop-out rate, 106 per group (i.e. 212 total) needed to be included.

The ‘recommended algorithm’ and ‘historic control’ cohorts were expected to be sufficiently comparable at baseline. However, matching was carried out between the two groups (24). The SAS PROC LOGISTIC was used to calculate the predicted probability of the dependent variable - the Propensity Score - for each observation in the data set. This single score (between 0 and 1) represents the relationship between multiple characteristics (i.e., the following baseline variables: age, sex, and comorbidities) and the dependent variable (i.e., the treatment group) as a single characteristic. Then the propensity score represents the predicted probability of receiving treatment. Using the SAS %MACRO OneToManyMTCH, the 108 ‘recommended algorithm’ individuals were matched to 108 ‘control’ subjects with the closest propensity score. Moreover, to verify the robustness of the above-described propensity score method, a further exploratory approach was performed by using the ‘teffects iptw’ STATA command to estimate the average treatment effect from observational data by inverse probability treatment weighting (IPTW), including 3368 patients in the control ORIGIN database.

Continuous variables were analyzed through descriptive statistics and reported as mean (SD) or median [IQR], as appropriate. Within-group changes with respect to baseline were analyzed with paired t-test or Wilcoxon signed-rank test, as appropriate. To determine the proportion of patients who required hospitalization a Log-rank test was used.

The cumulative costs for hospitalization in the two cohorts were the arithmetic sum of the direct cost of stay in an ordinary ward, sub-intensive care unit and intensive care unit for the entire period of hospitalization. In particular, in each cohort the total number of days that all patients spent in each of the three units of the hospital was multiplied by the corresponding estimated direct cost of stay per day (i.e., € 427, € 582, and € 1,278 per stay in an ordinary ward, and sub-intensive and intensive care units, respectively). Then the cumulative costs were calculated as the sum of the overall costs of stay in the three units. The direct cost per day was derived from data from a study on the management of COVID-19 patients admitted to hospital (Azienda Ospedaliera Nazionale SS. Antonio e Biagio e Cesare Arrigo, Alessandria, Italy) and the resources employed, performed by the Associazione Italiana Ingegneri Gestionali in Sanità (Castellanza, Varese, Italy) and presented at the LIUC Business School (Castellanza, Varese Italy) (http://www.liucbs.it – Webinar COVID, 8 July 2020).

All analyses were performed using SAS 9.4 (SAS Institute Inc, Cary, NC) and Stata 15 (StataCorp, College Station, TX). For the primary outcome a p-value of 0.05 was considered to determine statistical significance. For the six secondary outcomes a Bonferroni-adjustment for multiple tests was employed and a p-value of 0.0083 was used (25).

### 2.4 Ethical aspects

The COVER 2 study was approved by the Ethical Committee of Insubria (Varese, Italy; 27 July 2021) and registered at ClinicalTrials.gov (NCT04854824). In COVER 2, participants in the ‘recommended algorithm’ cohort provided written informed consent to their family doctors at enrolment. Subjects in the ‘control’ cohort (from the ORIGIN database) signed a consent form to participate in the ORIGIN study, which also explicitly included consent to use their data for future studies, such as COVER 2.

## 3. Results

### 3.1 Participants

Eight family doctors reported treating 108 consenting patients with early COVID-19 symptoms at home between January 2021 and May 2021, according to the proposed recommended algorithm (17). All individuals in this ‘recommendation’ cohort, had positive nasopharyngeal swabs, confirming SARS-CoV-2 infection. In 103 of 108 matched subjects identified in the ORIGIN dataset (‘control’ cohort) the onset of COVID-19 symptoms occurred between late February and July 2020, and in the other 5 participants between September 2020 and January 2021. SARS-CoV-2 infection was confirmed in all cases by nasopharyngeal swabs or serology tests. These individuals were treated at home by their family physicians with drug regimens not necessarily guided by those proposed in the recommendation algorithm. The cohorts were comparable in terms of mean age and age range, with most subjects aged between 41 and 65 (Table 1). Females were more prevalent in both cohorts (57.4% and 64.8%). The concomitant diseases were well distributed between the two groups, except for overweight/obesity, which were reported in a few more individuals in the ‘control’ cohort. The most common symptoms at the onset of illness were fever (70.4% vs 72.2%) and tiredness (68.5% vs 76.9%), followed by cough (60.2% vs 48.2%), and myalgia (48.2% vs 53.7%) in both the ‘recommendation’ and ‘control’ cohorts (Table 1). More individuals in the ‘recommended algorithm’ cohort had arthralgia (30.6% vs 3.7%, P=0.001), while ageusia was significantly more frequent in the ‘control’ cohort (38.9% vs 55.6%, P=0.020). The distribution of dyspnea was similar between the two groups (25.9% vs 31.5%, P=0.452).

**Table 1.** Demographic and early symptoms associated with COVID-19 illness in the two treatment cohorts.

|  | **Overall**<br>**(*n*=216)** | **Recommended**<br>**treatment**<br>**cohort**<br>**(*n*=108)** | **Control**<br>**cohort**<br>**(*n*=108)** | **P value** |
| --- | --- | --- | --- | --- |
| **Demographic characteristics** |  |  |  |  |
| Age, years |  |  |  |  |
| 18-40 | 43 (19.90) | 23 (21.30) | 20 (18.52) | 0.968 |
| 41-65 | 127 (58.80) | 63 (58.34) | 64 (59.26) |  |
| 66-75 | 23 (10.65) | 11 (10.18) | 12 (11.11) |  |
| >75 | 23 (10.65) | 11 (10.18) | 12 (11.11) |  |
| Mean age ± SD | 53.3±15.4 | 53.1±15.8 | 53.5±15.1 | 0.847 |
| Males, *n* (%) | 84 (38.89) | 46 (42.59) | 38 (35.18) | 0.329 |
| **Comorbidities, *n* (%)** |  |  |  |  |
| Cardiovascular disease | 19 (8.80) | 8 (7.41) | 11 (10.18) | 0.652 |
| Hypertension | 51 (23.61) | 23 (21.30) | 28 (25.93) | 0.522 |
| Diabetes mellitus | 5 (1.85) | 1 (0.93) | 4 (3.70) | 0.369 |
| Overweight/Obesity | 33 (15.28) | 11 (10.18) | 22 (20.37) | 0.057 |
| Chronic kidney disease | 1 (0.46) | 1 (0.93) | 0 (0) | 1.000 |
| **Early symptoms, *n* (%)** |  |  |  |  |
| Fever | 154 (71.30) | 76 (70.37) | 78 (72.22) | 0.880 |
| Myalgia | 110 (50.92) | 52 (48.15) | 58 (53.70) | 0.496 |
| Arthralgia | 37 (17.13) | 33 (30.55) | 4 (3.70) | 0.001 |
| Tiredness/exhaustion | 157 (72.68) | 74 (68.52) | 83 (76.85) | 0.222 |
| Dyspnea | 62 (28.70) | 28 (25.93) | 34 (31.48) | 0.452 |
| Chest pain | 32 (14.81) | 14 (12.96) | 18 (16.67) | 0.566 |
| Headache | 87 (40.28) | 46 (42.59) | 41 (37.96) | 0.579 |
| Lack of appetite | 64 (29.63) | 26 (24.07) | 38 (35.18) | 0.101 |
| Cough | 117 (54.17) | 65 (60.18) | 52 (48.15) | 0.101 |
| Sore throat | 57 (26.39) | 35 (32.41) | 22 (20.37) | 0.063 |
| Rhinitis | 59 (27.31) | 31 (28.70) | 28 (25.93) | 0.760 |
| Vomiting/nausea | 33 (15.28) | 13 (12.04) | 20 (18.52) | 0.256 |
| Diarrhea | 38 (17.59) | 16 (14.81) | 22 (20.37) | 0.372 |
| Red eyes | 22 (10.18) | 7 (6.48) | 15 (13.89) | 0.114 |
| Vertigo | 11 (5.09) | 10 (9.26) | 1 (0.93) | 0.010 |
| Sicca syndrome | 1 (0.46) | 1 (0.93) | 0 (0) | 1.000 |
| Anosmia | 88 (40.74) | 37 (34.26) | 51 (47.22) | 0.071 |
| Ageusia | 102 (47.22) | 42 (38.89) | 60 (55.55) | 0.020 |

Data are numbers (percentages). Between-group differences were assessed by Fisher's exact test.

### 3.2 Primary outcome

One of the 108 patients (0.9%) in the ‘recommended’ cohort was hospitalized, compared to 12 of the 108 patients (11.1%) in the ‘control’ cohort (Figure 1). The event rate was significantly lower in the ‘recommended’ than in the ‘control’ group (survival analysis for clustered data, P=0.0136) (Figure 1). The patient in the ‘recommended’ cohort was admitted to hospital due to dyspnea secondary to interstitial pneumonia (Table 2). This was the same reason for the hospitalization of all patients in the ‘control’ cohort, except for one who was admitted with dyspnea due to documented pulmonary thromboembolism (Table 2).

**Table 2.** Clinical course of hospitalized patients in the two cohorts.

| Cohort | Reason for hospital admission | Hospitalisation (days) | Oxygen therapy* (yes/no) | CPAP (yes/no) | CPAP (days) | Mechanical ventilation (yes/no) | Mechanical ventilation (days) | ICU admission (yes/no) | ICU admission (days) | Sequelae at discharge (yes/no) |
| --- | --- | --- | --- | --- | --- | --- | --- | --- | --- | --- |
| *Control* |  |  |  |  |  |  |  |  |  |  |
| Control | Dyspnea (interstitial pneumonia) | 12 | Yes | No | - | No | - | No | - | No |
| Control | Dyspnea (interstitial pneumonia) | 26 | Yes | No | - | No | - | No | - | No |
| Control | Dyspnea (interstitial pneumonia) | 4 | Yes | No | - | No | - | No | - | No |
| Control | Dyspnea (interstitial pneumonia) | 12 | Yes | No | - | No | - | No | - | No |
| Control | Dyspnea (interstitial pneumonia) | 4 | Yes | No | - | No | - | No | - | No |
| Control | Dyspnea (interstitial pneumonia) | 13 | Yes | No | - | No | - | No | - | No |
| Control | Dyspnea (interstitial pneumonia) | 17 | Yes | No | - | No | - | No | - | No |
| Control | Dyspnea (interstitial pneumonia) and epigastralgia | 6 | No | No | - | No | - | No | - | No |
| Control | Dyspnea (interstitial pneumonia) | 10 | Yes | No | - | No | - | No | - | No |
| Control | Dyspnea (interstitial pneumonia) and epigastralgia | 9 | Yes | No | - | No | - | No | - | No |
| Control | Dyspnea<br>(interstitial<br>pneumonia) and<br>gastrointestinal<br>symptoms | 8 | Yes | No | - | No | - | No | - | No |
| Control | Dyspnea<br>(Pulmonary<br>thromboembolism) | 20 | No | No | - | No | - | No | - | No |
| *'Recommended'* |  |  |  |  |  |  |  |  |  |  |
| 'Recommended' | Dyspnea<br>(interstitial<br>pneumonia) | 19 | Yes | Yes | 6 | No | - | No | - | No |

\*Conventional oxygen therapy (oxygen delivered by nasal tube, nasal cannula, or face mask). ° CPAP, continuous positive airway pressure; ICU, intensive care unit.

**Figure 1.**
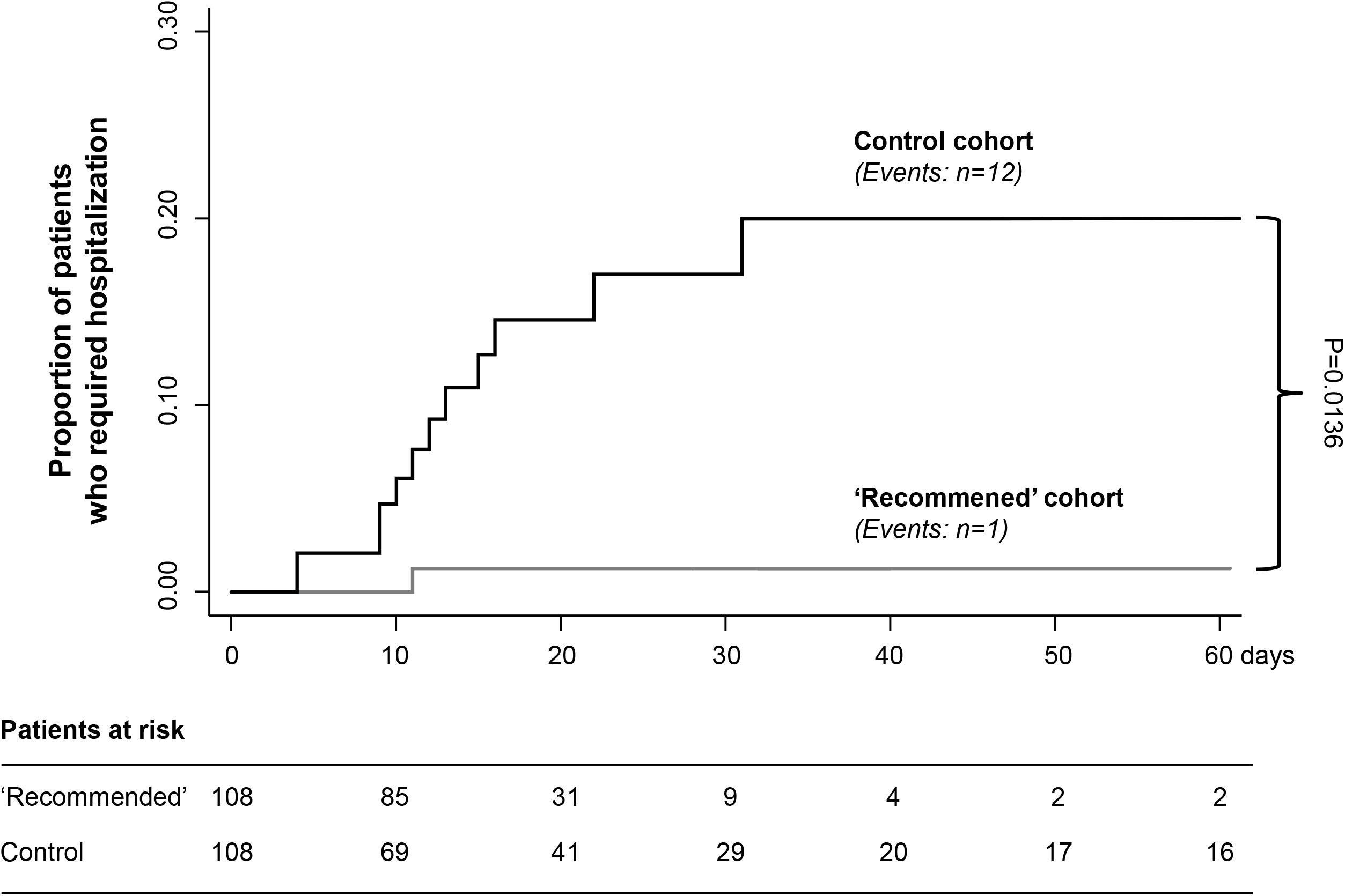
Kaplan-Meier curves for the primary endpoint of hospital admission. Kaplan-Meier curves show the proportion of patients who required hospitalization in the two treatment cohorts. Grey line, ‘recommended algorithm’ treatment cohort; black line, ‘control’ cohort. P-value for treatment comparison was assessed by survival analysis for clustered data.

To confirm these findings, explorative analysis was performed using the inverse probability weighting (IPTW) method, including 3368 patients in the control ORIGIN database. We found that the hospitalization rate in the ‘recommended algorithm’ cohort was significantly lower than in the ‘control’ cohort (-0.059; 95% CI, -0.077 to -0.041; P<0.0001).

### 3.3 Secondary outcomes

Seventy-four of 108 ‘recommended’ cohort patients were treated with a relatively selective COX-2 inhibitor, such as nimesulide or celecoxib, while 15 patients were given aspirin (Table 3). Non-adherence to the recommended anti-inflammatory regimen was 24.07%, since 26 patients were prescribed other NSAIDs (ketoprofen, ibuprofen or paracetamol). In the ‘recommended’ cohort, anti-inflammatory treatment with NSAIDs was prescribed by family physicians within a mean (± SD) of 1.7 ± 3.3 days after the onset of symptoms, except for paracetamol that was self-administered by the patients before contacting the doctor. At variance, in the ‘control’ cohort only few patients received relatively selective COX-2 inhibitors (n=4) or aspirin (n=5) (Table 3). Notably, in this cohort most patients were given paracetamol (n=74), and the remaining ketoprofen or ibuprofen.

**Table 3.** Treatment at home in the two study cohorts.

|  | **Recommended**<br>**treatment cohort**<br>**(n=108)** | **Control cohort**<br>**(n=108)** | **P value** |
| --- | --- | --- | --- |
| **Relatively selective COX-2 inhibitors** | 74 (68.52) | 4 (3.70) | P<0.001 |
| Nimesulide | 36/74 (48.65) | 1/4 (25.00) |  |
| Morniflumate | 0 (0) | 2 (50.00) |  |
| Celecoxib | 38/74 (51.35) | 0/ (0) |  |
| Etoricoxib | 0/ (0) | 1/ (25.00) |  |
| **Other NSAIDs** | 34 (31.48) | 82 (75.93) | P<0.001 |
| Aspirin | 15/34 (44.12) | 5/82 (6.10) |  |
| Ketoprofen | 7/34 (20.59) | 4/82 (4.88) |  |
| Ibuprofen | 10/34 (29.41) | 12/82 (14.63) |  |
| Indomethacin | 0/ (0) | 0/ (0) |  |
| Paracetamol | 9/34 (26.47) | 74/82 (90.24) |  |
| **Corticosteroids** | 28 (25.93) | 7 (6.48) | P<0.001 |
| **Anticoagulants** | 3 (2.78) | 2 (1.85) | P=1.000 |
| **Antibiotics** | 41 (37.96) | 26 (24.07) | P=0.039 |
| *Azithromycin* | 20/41 (48.78) | 7/26 (26.92) |  |
| *Amoxicillin and clavulanic acid* | 0/41 (0) | 3/26 (11.54) |  |
| **Need of oxygen\*** | 10 (9.26) | 2 (1.85) | P=0.033 |

Data are n/N (percentages). COX-2, cyclooxygenase-2; NSAIDs, nonsteroidal anti-inflammatory drugs. \* Need for oxygen therapy at home. Between-group differences were assessed by Fisher's exact test.

Corticosteroids were prescribed to 26% and 6.5% of patients in the ‘recommended’ and ‘control’ cohorts, respectively (P<0.001) (Table 3). A median of 7 [IQR: 5-8.5] days elapsed between starting NSAID and corticosteroid prescriptions in the ‘recommended’ group. More patients were treated with antibiotics in the ‘recommended’ than in the ‘control’ cohort (P=0.039), while anticoagulants were prescribed in very few cases in both groups (Table 3). Ten patients in the ‘recommended’ cohort and two in the ‘control’ cohort required oxygen supply at home due to decreasing oxygen saturation or following a first episode of dyspnea or wheezing (P=0.033) (Table 3).

Almost all patients achieved resolution of the major symptoms (i.e., complete remission), and the event rate was numerically - but not significantly - higher in the recommended than in the ‘control’ cohort between the two cohorts (P=0.332) (Table 4). On the other hand, the proportion of patients with persistent other symptoms, such as anosmia, ageusia/dysgeusia, lack of appetite and fatigue was significantly lower in the ‘recommended’ than in the ‘control’ cohort (20.4% vs 63.9%, respectively; P<0.001) (Table 4). This difference was shown in the subgroups of patients with these symptoms persisting for 30 to 60 days or more than 60 days (Table 4).

**Table 4.** Major secondary outcomes.

|  | **Recommended treatment cohort**<br>(*n*=108) | **Control cohort**<br>(*n*=108) | **Nominal P value** |
| --- | --- | --- | --- |
| Time from symptoms onset and start of anti-inflammatory therapy (days) | 1.7 ± 3.3 | - | - |
| Rate of resolution of major symptoms* | 105/108 (97.2) | 101/108 (93.5) | P=0.332 |
| Rate of persistence of other symptoms<sup>°</sup> | 22/108 (20.4) | 69/108 (63.9) | P<0.001** |
| Persistence of other symptoms (days) |  |  |  |
| < 30 | 6/22 (27.3) | 13/69 (18.8) | P=0.385 |
| 30-60 | 8/22 (36.4) | 6/69 (8.7) | P=0.0043** |
| >60 | 8/22 (36.4) | 50/69 (72.5) | P=0.0043** |

Data are n/N (percentages) or mean ± SD, as appropriate. \* defined as complete recovery from major symptoms, i.e., no fever, SpO<sub>2</sub>>94% and/or no dyspnea, no cough, no rhinitis, no pain (myalgia, arthralgia, chest pain, headache, sore throat), no vertigo, no nausea, vomiting or diarrhea, no sicca syndrome or red eyes; <sup>°</sup> defined as recovery from major COVID-19 symptoms, but persistence of symptoms such as anosmia, ageusia/dysgeusia, lack of appetite, fatigue. \*\* Significant after Bonferroni adjustment for multiple tests.

The single patient in the ‘recommended’ cohort who was hospitalized was discharged after 19 days, compared to 12±7 (range, 4 to 26) days in the 12 patients in the ‘control’ cohort. The cumulative length of hospital stays in the latter cohort reached 141 days (Table 2). At variance with the patient in the ‘recommended’ cohort who spent 6 days in a sub-intensive care unit and 13 days in the ordinary unit, none of the patients in the ‘control’ cohort required admission to sub-intensive care units or an ICU, and all were managed in the ordinary hospital units (Table 2). Thus, cumulative hospitalization costs were € 9.058 and € 60.316 in the ‘recommended’ and ‘control’ cohorts, respectively (Figure 2). Only 9.8 (95% CI: 6.1 to 25.1) patients needed to be treated with the home therapy algorithm to prevent one hospitalization event.

**Figure 2.**
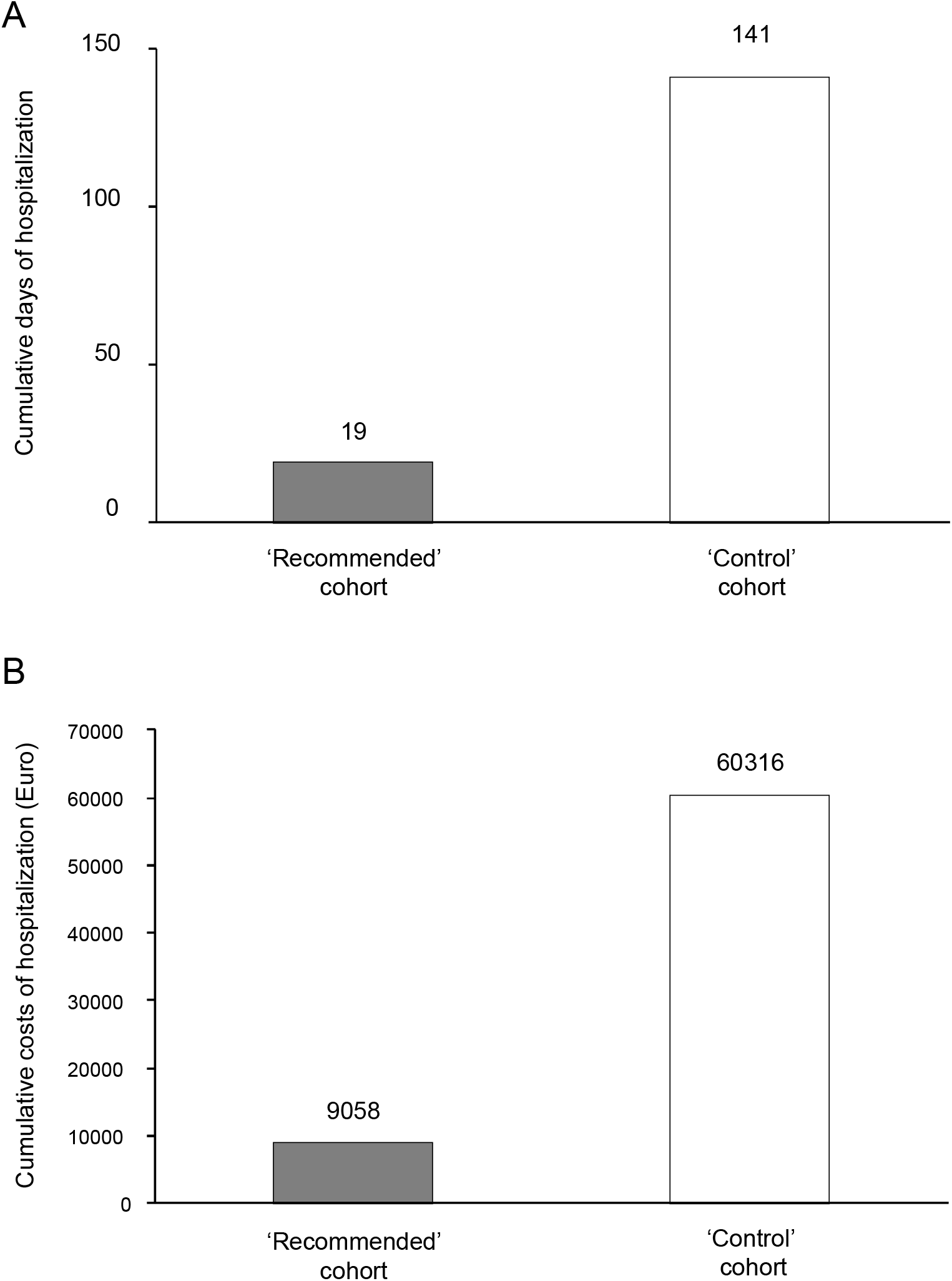
Cumulative length of hospital stay and related costs in the two study cohorts. Cumulative days of hospitalization **(A)** and cumulative costs for hospital stay **(B)** in the ‘recommended’ treatment cohort and in the ‘control’ cohort. Grey columns, ‘recommended’ treatment cohort; white columns, ‘control cohort’.

Regarding hospital admission, a sensitivity analysis was also performed after excluding patients who spontaneously initiated paracetamol before the family doctor prescriptions in the ‘recommended’ cohort and the related matched patients in the ‘control’ cohort. Similarly to the intention-to-treat analysis, only 1 of the 99 patients (1.01%) in the ‘recommended’ cohort required hospital admission, compared to 10 of the 99 patients (10.1%) in the ‘control’ cohort. The event rate was still significantly lower in the ‘recommended’ than in the ‘control’ cohort (survival analysis for clustered data, P=0.0193).

## 4. dISCUSSION

In this observational matched-cohort study we found that COVID-19 patients being treated at home early after the onset of symptoms by their family physicians according to the proposed recommendation algorithm almost completely prevented the need for hospitalization due to severe worsening of the illness (primary outcome of the study), compared to patients in the ‘control’ cohort, who were treated at home according to their family doctor’s judgment. This resulted in an over 85% reduction in the length of hospital stays, which translated into a similar percentage of lowered related treatment costs. Thus, the cost-effectiveness of the home recommendation treatment algorithm was remarkable, considering that in the two cohorts the early symptoms were comparable. In line with this observation, only 9.8 patients needed to be treated to prevent one hospitalization event. These findings, achieved in a larger number of COVID-19 patients, further corroborate the results of our previous matched-cohort study regarding the lower risk of hospital admission in patients treated at home at the onset of illness according to the recommendation algorithm (20), than with other regimens. Similarly, the rate of resolution of major COVID-19 symptoms, including fever, myalgias/arthralgias, headache and cough, was numerically higher in the ‘recommended’ algorithm than in the ‘control’ cohort. Moreover, other symptoms, such as anosmia or ageusia, or fatigue, ceased more frequently and persisted for a shorter period in the ‘recommended’ than in the ‘control’ cohort. Together, these observations suggest that the two regimens targeting early symptoms, not the virus, affected COVID-19 disease phenotype in different ways, which translated into a remarkable decreased need for hospitalization in patients treated according to the ‘recommended’ algorithm. Moreover, the lower hospitalization rate in this cohort cannot be attributed to limited access to hospitals, since patients in the ‘recommended’ regimen group became ill during the third wave of the pandemic (from January 2021), when hospital (human and technical) resources were brought close to but did not reach the limit at which they would have been forced to deny hospital admission of those with severe COVID-19. This was not the case for the ‘control’ cohort, in which most patients reported symptoms during the first stage of the COVID-19 outbreak, when hospitals were under huge pressure, which may have resulted in postponed or denied hospitalization for some patients in need. Thus, finding that there was a remarkably higher hospitalization rate in the ‘control’ cohort provided additional evidence of the protective effect of the proposed treatment algorithm against hospitalization because of worsening COVID-19 symptoms.

Our recommendation treatment algorithm (17) is based on the idea that it is critical to intervene at home very early on during the onset of mild/moderate symptoms to avoid progression toward severe COVID-19, which would eventually require hospital admission. Indeed, after the initial exposure to SARS-CoV-2, patients typically develop symptoms that indicate an inflammatory process within 5 to 6 days on average (15,16), and pro-inflammatory mediators, in particular cytokines, seem to be integral to the initiation, intensification, propagation and worsening of tissue morbidity related to COVID-19 (16,19,26). Therefore, our recommended treatment algorithm moved from this pathophysiologic rationale of early COVID-19 events, and focused on the initial use of NSAIDs, which has been shown to reduce pro-inflammatory cytokine levels (18). NSAIDs inhibit the cyclooxygenase activity of prostaglandin H synthase 1 and 2, also named COX-1 and COX-2 (27). Relatively selective COX-2 inhibitors (e.g., celecoxib, etoricoxib) (27) may reduce pro-inflammatory cytokine levels, as shown in mice with influenza A infection (TNF-α, G-CSF, and IL-6) (28) and in hospitalized COVID-19 patients (IL-6) (19,29). The overlap in COX-2 selectivity between coxibs and the more traditional NSAID nimesulide (27) was the rationale for recommending these drugs for the treatment of early COVID-19 at home, unless contraindicated. Adherence to this recommendation was very high (75.3%) in the ‘recommended’ algorithm cohort. On the other hand, in the ‘control’ cohort very few patients were treated with a COX-2 inhibitor, and most received paracetamol. However, this drug, considered an alternative for addressing the symptoms of COVID-19 in the early stages (14), has negligible anti-inflammatory effect (30), in addition to being capable of inducing or worsening glutathione consumption (31,32). Given the anti-oxidant property of glutathione, it has recently been hypothesized that paracetamol might even exacerbate COVID-19 (31,32).

Physicians may be reluctant to use NSAIDs, including relatively selective COX-2 inhibitors, due to the known risk of cardiovascular events (33) and the hepatotoxicity of nimesulide, which is admittedly very low when the drug is prescribed at the recommended daily dose and time of administration (34). On the other hand, in a large cohort of over 4200 patients admitted to the hospital who had taken NSAIDs within the 2 weeks preceding hospital admission, the use of these drugs was not associated with higher mortality or increased severity of COVID-19, as compared to a matched group of NSAID non-users (35). Moreover, another study provided no indication that harm was induced by NSAIDs, as demonstrated by the lack of increased risk of poorer outcomes in COVID-19 patients given NSAIDs compared with those treated with paracetamol, or NSAID non-users (36). None of the patients in the ‘recommended algorithm’ cohort developed toxicity related to or possibly related to the use of celecoxib or nimesulide. This is in line with the fact that few patients in this cohort received aspirin, which the recommendations propose as alternative therapy when contraindications to celecoxib or nimesulide are highlighted by physicians. Notably, there is evidence that aspirin may reduce plasma levels of pro-inflammatory cytokines (37), and lower the risk of in-hospital mortality in a large cohort of patients hospitalized with COVID-19 (38), supporting the use of this drug in the early stages of COVID-19 at home when needed. In the future, other NSAIDs, such as indomethacin, which also lowers IL-6 in SARS-CoV-2 patients (39), could be proposed as an alternative treatment for early COVID-19 symptoms at home, as anticipated by a recent small Indian study (40).

The same pharmacologic rationale was adopted for the recommendation of the use of corticosteroids, known to exert their anti-inflammatory effects mainly by inhibiting pro-inflammatory genes that encode for cytokines and chemokines (41). Our proposal clearly suggests only starting corticosteroid several days after the onset of symptoms if fever or musculoskeletal pain persist despite NSAIDs or when oxygen saturation significantly declines. According to this, in the ‘recommended algorithm’ cohort, corticosteroids were administered only after a median of 7 days after the onset of symptoms and when they fulfilled the proposed criteria for starting this class of drugs, not necessarily limited to patients in need of oxygen supply. This might explain the discrepancy between the number of patients treated with corticosteroids (n=28) and those given oxygen therapy (n=10) in the ‘recommended’ cohort. Despite concerns about the use of corticosteroids in COVID-19 patients due to the risk of complications and the possible persistence of the virus in the host (42,43), no side effects related to the use of these drugs were reported in patients of the ‘recommended’ cohort. Based on the large RECOVERY trial (10), WHO recommended systemic corticosteroids only in hospitalized patients with severe COVID-19 who require respiratory support (44). However, there is also some evidence of the benefit of corticosteroids during the early phase of the illness (45,46), recently corroborated by findings of randomized controlled trials with inhaled corticosteroids in the community (47,48). The administration of inhaled budesonide within 7 days of the onset of mild COVID-19 symptoms markedly reduced the risk of hospitalization compared to patients receiving the usual care, results which are similar to those achieved in our ‘recommended algorithm’ cohort. Interestingly, the recommendations of the Italian Ministry of Health for the management of COVID-19 patients at home have recently been updated (14) to include corticosteroids for the treatment of early COVID-19 symptoms according to criteria very similar to those proposed in our recommendation algorithm (17).

Despite being recommended by the algorithm, especially for those bedridden or with high D-dimer levels, only a small number of COVID-19 patients in the ‘recommendation’ cohort received a prophylactic dose of LMW heparin. None of them had side effects. Actually, COVID-19 is characterized by dysregulation of the coagulation system and fibrinolysis that can promote micro- and macro-vascular thrombosis, as well as venous thromboembolic complications, which are sometimes life-threatening (16,49,50). Even guidelines (14) suggest that LMW heparin be used at a prophylactic dosage in COVID-19 patients at home in particular instances. Nonetheless, a recent study involving 2219 noncritically ill, hospitalized COVID-19 patients reported that therapeutic-dose anticoagulation with heparin increased the probability of in hospital survival compared with standard care thromboprophylaxis, regardless of the patient’s baseline D-dimer levels (51). This finding creates the possibility of studying an initial strategy of therapeutic versus prophylactic anticoagulation with LMW heparin in COVID-19 patients with moderate symptoms who are being treated at home as well.

Similarly, the recommendation for antibiotic treatment was just in case of suspected bacterial pneumonia or suspected secondary bacterial upper respiratory infections, not on a routine basis, which is in line with the UK NICE COVID-19 guidelines for managing patients at home (13). According to these indications, family doctors in the ‘recommended algorithm’ cohort used antibiotics in 37% of their COVID-19 patients. This is not surprising, considering that in a systematic review on hospitalized COVID-19 patients, 1450 of 2010 individuals (72%) were treated with antibiotics, despite only 8% presenting with evidence of bacterial coinfection (52). Nonetheless, the risk of developing antimicrobial resistance should invite caution regarding the indiscriminate use of antibiotics.

The non-randomized design is a major limitation of the study, which is observational in nature. Nonetheless, comparative analysis of patient cohorts in everyday clinical practice with adjustments for possible confounding biases may offer a suitable alternative to the recommended clinical trials to evaluate the effectiveness of different therapeutic regimens (53,54). Moreover, the matched-cohort study protocol with a statistical plan was predefined and the analyses were performed accordingly. There is the additional limitation that the collection of outcome information in the ‘control’ cohort was through interviews and questionnaires related to events that happened before the survey. This was not the case for the ‘recommended algorithm’ cohort, where data were gathered by family doctors. However, in both cohorts the date of hospital admission (primary outcome) and data about the course of hospitalization were well documented in the hospital discharge letter. Moreover, further evidence of the observed difference between the hospital admission rates for the two cohorts is offered by the results of the additional explorative analysis of 3368 patients in the control ORIGIN database, which confirmed a significantly lower rate of hospitalization in the ‘recommended algorithm’ than in the ‘control’ group.

On the other hand, the COVER 2 study formally tested outcomes for COVID-19 patients managed by their family physicians according to a therapy recommendation algorithm that targets early symptoms, based on the pathophysiology of the illness and the related pharmacologic rationale. This is a strength of the COVER 2 study, since none of the recently proposed recommendations on how to treat COVID-19 patients for family doctors in the community have been formally evaluated on whether they can limit the progression of mild/moderate symptoms at the onset of the disease to the need for hospital admission.

In conclusion, we have documented that simple, reasoned treatments for the early-phase symptoms of COVID-19 at home, collected in a recommendation algorithm for family doctors, are beneficial in clinical practice, since they may avoid or limit deterioration of the disease to the point of hospitalization, in addition to having public health implications. Our findings also have important implications for patient quality of life, since adopting the treatment recommendation approach reduced the rate and shortened the duration of symptoms, such as loss of taste or smell, and fatigue, which might otherwise persist for several months (55). Future randomized studies will be required for the consolidation of these observational findings on the potential benefit of the proposed treatment recommendation algorithm.

## Data Availability

Sharing individual participant data with third parties was not specifically included in the informed consent form of the study, and unrestricted diffusion of such data may pose a potential threat of revealing the identities of participants, as permanent data anonymization was not carried out (patient records were instead de-identified per protocol during the data retention process). To minimize this risk, individual participant data that underlie the results reported in this article will be available after three months and for up to five years following article publication. The researchers shall submit a methodologically sound proposal to Dr. Annalisa Perna, Head of the Laboratory of Biostatistics of the Department of Renal Medicine of the Istituto di Ricerche Farmacologiche Mario Negri IRCCS. To gain access, data requestors will need to sign a data access agreement and obtain the approval of the local ethics committee.

## Author Contributions

GR, FS and PR had the original idea; NP and GR wrote the draft version of the manuscript; EC, SP, CM, EP, MVP, GP, UC, FS contributed to patient identification; NR helped with data collection and management; AP, TP performed the statistical analyses; NP, PR, GR developed the final version of the manuscript, all authors critically revised the final version. GR and NP took responsibility for the submission for publication. No medical writer was involved.

## Ethics statement

The COVER 2 study has been approved by the Ethical Committee of Insubria (Varese, Italy; 27 July 2021) and registered at the ClinicalTrials.gov (NCT04854824). In COVER 2, participants in the ‘recommended algorithm’ cohort provided written informed consent to their family doctors at enrolment. Subjects in the ‘control’ cohort (from the ORIGIN database) signed a consent form to participate in the ORIGIN study, which also explicitly included consent to use their data for future studies, such as COVER 2.

## Data Availability Statement

Sharing individual participant data with third parties was not specifically included in the informed consent form of the study, and unrestricted diffusion of such data may pose a potential threat of revealing participants’ identities, as permanent data anonymization was not carried out (patient records were instead de-identified per protocol during the data retention process). To minimize this risk, individual participant data that underlie the results reported in this article will be available after three months and for up to five years following article publication. The researchers shall submit a methodologically sound proposal to Dr. Annalisa Perna, Head of the Laboratory of Biostatistics of the Department of Renal Medicine of the Istituto di Ricerche Farmacologiche Mario Negri IRCCS. To gain access, data requestors will need to sign a data access agreement and obtain the approval of the local ethics committee.

