## Supplemental Material for "A HOME-TREATMENT ALGORITHM BASED ON ANTI-INFLAMMATORY DRUGS TO PREVENT HOSPITALIZATION OF PATIENTS WITH EARLY COVID-19: *A MATCHED-COHORT STUDY (COVER 2)*"

**SUPPLEMENTARY MATERIAL - COVER 2 study**

**Summary of proposed recommendations for treatment**

### **of COVID-19 patients at home**

Recommended treatments should start immediately when COVID-19 early symptoms appear without waiting results of a nasopharyngeal swab, if any. The recommended drugs can be used unless contraindicated according to the summary of product characteristics.

I. Non-steroidal anti-inflammatory drugs (NSAIDs)

*Relatively selective COX-2 inhibitors* ^§#^ (*for myalgias and/or arthralgias or other painful symptoms*)

*^§^ based on the ratio of concentrations of the various NSAIDs required to inhibit the activity of COX-1 and COX-2 by 50 percent (IC_50_) in assays of whole blood*

*^#^unless contraindicated*

*Nimesulide **

100 mg b.i.d p.o, after a meal, for a maximum of 12 days.

*Or*

*Celecoxib **

Initial oral dose of 400 mg, followed by a second dose of 200 mg on the first day of therapy. In the following days, up to a maximum of 400 mg (200 mg twice a day) should be given as needed for a maximum of 12 days

** Should the patient have fever (≥ 37.3 °C) or develop laboratory signs of hepatotoxicity associated with nimesulide or there are contraindications to celecoxib, these drugs* *should be substituted with aspirin* *(a COX-1 and COX-2 inhibitor) (500 mg twice a day p.o. - after a meal). These treatments should be associated with a proton pump inhibitor (e.g. lansoprazole - 30 mg/day; or omeprazole - 20 mg/day; or pantoprazole - 20 mg/day).*

*After approximately 3 days from the onset of symptoms (or more days are elapsed and the physician sees the patient for the first time), a series of hematochemical tests should be performed (blood cell count, D-dimer, CRP, creatinine, fasting blood glucose, ALT). Should inflammatory indexes (CRP, neutrophil count), ALT, and D-dimer be in the normal range, nimesulide****/****celecoxib (or aspirin) treatment will continue.*

II. Corticosteroids*

Dexamethasone (*for persistent fever or musculoskeletal pain or when few days later hematochemical tests were repeated and even mild increase of inflammatory indexes - CRP, neutrophil count - are documented, or cough and oxygen saturation (SpO_2_) <94-92% occur)*

8 mg p.o for 3 days, then tapered to 4 mg for a further 3 days, and then to 2 mg for 3 days. That makes 42 mg dexamethasone total over 9 days.

**Duration of corticosteroid treatment also depends on the clinical evolution of the disease*

III. Anticoagulants

*Low-molecular weight (LMW) heparin** *(when the hematochemical tests show even a mild increase of D-dimer or for thromboembolism prophylaxis for bedridden patients)*

Enoxaparin, at the prophylactic daily dose of 4000 U.I subcutaneously - i.e. 40 mg enoxaparin. Treatment recommended for at least 7-14 days, independently of the patient recovering mobility.

**unless contraindicated (e.g., ongoing bleeding or platelet count <25 x 10^9^/L)*

IV. Oxygen therapy

Gentle oxygen supply in the early phase of the disease, possibly before pulmonary symptoms manifest, in the presence of progressively decreasing oxygen saturation – as indicated by oximeter – or following a first episode of dyspnoea or wheezing.

Conventional oxygen therapy is suggested when the respiratory rate is >14/min and oxygen saturation (SpO_2_) *<94-92%,* but is required with SpO_2_ <90% at room air. With liquid oxygen, start with 8-10 litre/min and monitor SpO_2_ every 3-4 hours. Titrate oxygen flow rate to reach target SpO_2_ *>94%.* Then the rate of oxygen administration can be reduced to 4-5 litre/min (but continue SpO_2_ monitoring every 3-4 hours). With gaseous O_2,_ start with 2.5-3.0 litre/min, but monitor SpO_2_ more frequently than with liquid oxygen, and titrate flow rates to reach target SpO_2_ *>94%.* Should patients be poorly responsive to high O_2_ administration, consider hospitalisation, if feasible.

V. Antibiotics

*Azithromycin** *(with bacterial pneumonia or suspected secondary bacterial upper respiratory tract infections, or in particularly fragile patients, or when hematochemical inflammatory indexes (CRP, neutrophil count) are markedly altered)*

500 mg/day p.o for 6-10 days depending on the clinical judgement

* *Should the patient be at risk of or with a history of cardiac arrhythmia or present other contraindications, cefixime (400 mg/day p.o for 6-10 days) or amoxicillin/clavulanic acid (1gr three times a day for 6-10 days)* *can be considered as alternative to azithromycin.*

**COVER 2 Study Organization**

Members of the COVER 2 Study Organization were as follows (all in Italy): *Chief Investigator* - Giuseppe Remuzzi (Bergamo); *Study coordinators* - Norberto Perico, Fredy Suter (Bergamo); *Coordinating Centre* – Istituto di Ricerche Farmacologiche Mario Negri IRCCS, Centro di Ricerche Cliniche per le Malattie Rare *Aldo e Cele Daccò*, Ranica (Bergamo); *Study investigators including patients* - Elena Consolaro (Varese), Chiara Moroni (Varese), Umberto Cantarelli (Teramo), Stefania Pedroni (Varese), Maria Vittoria Paganini (Varese), Elena Pastò (Varese), Grazia Pravettoni (Varese), Fredy Suter (Bergamo); *Data collection and processing* –Nadia Rubis, Davide Villa, Olimpia Diadei, Davide Martinetti, Matias Trillini (Bergamo); *Data Analysis* - Annalisa Perna, Tobia Peracchi (Bergamo); *Regulatory Affairs* - Paola Boccardo (Bergamo); *Finalization of the manuscript*: Norberto Perico, Piero Ruggenenti, Giuseppe Remuzzi (Bergamo).
